# Transcriptome-wide association implicates *HTRA1-AS1* in age-related macular degeneration risk

**DOI:** 10.64898/2025.12.05.25341732

**Authors:** Inti Pagnuco, Jacob Sampson, Andrew P Morris, Jamie M Ellingford

## Abstract

Age-related macular degeneration (AMD) is a leading cause of irreversible visual impairment and legal blindness that has significant genetic risk factors, including genetic variation within a locus on chromosome 10 encompassing several extensively studied protein-coding genes (*ARMS2, HTRA1* and *PLEKHA1*). Here, through transcriptome-wide association and fine-mapping approaches that integrate newly available high coverage transcriptome datasets for the human eye, we provide robust statistical association between elevated AMD-risk and genetic regulation of a long non-coding RNA gene within the chromosome 10 locus, *HTRA1-AS1*. Our findings show that increased expression of *HTRA1-AS1* elevates AMD-risk and provides a new paradigm for the study of biological drivers of AMD-risk and early disease pathogenesis.

## Main

Age-related macular degeneration (AMD) is characterised by progressive and irreversible loss of central detailed vision in adults over 55 [1, 2] and is predicted to impact over 280 million individuals by 2040 [3]. Genetic risk factors for AMD have been well established through genome-wide association studies (GWAS) [4-12], with over 60 genomic loci identified at genome-wide significance. The most commonly and strongly associated genetic risk loci for AMD map to regions on chromosome 1, including complement factor H (*CFH*) [4-6], and on chromosome 10, including age-related macular susceptibility 2 (*ARMS2*) [13, 14] and high temperature requirement A serine peptidase 1 (*HTRA1*) [9, 11]. The biological processes underpinning chromosome 1 and chromosome 10 AMD-risk are currently thought to be distinct [2]. However, there are discordant findings of the molecular mechanisms driving the *ARMS2/HTRA1* AMD association at the chromosome 10 locus [15], which is complicated by a region of high linkage-disequilibrium (LD) making it challenging to pinpoint credible independently associated variants through GWAS [16]. More recently the integration of co-localisation analysis with characterised expression quantitative trait loci (eQTL) for retinal tissue, and transcriptome-wide association studies (TWAS), have identified additional AMD-risk loci driven by changes in gene expression in the retina [17-19], but notably have not provided additional associations at the chromosome 10 risk locus.

In this study, we integrated newly available high coverage transcriptome datasets for the adult human retina [20] with recent large-scale AMD-GWAS findings [8, 12] using developments in TWAS and fine-mapping to further investigate AMD-risk mediated by genetic drivers of gene expression. Specifically, we built predictive gene models for retinal gene expression through the unified test for molecular signatures (UTMOST) statistical framework [21], using genotype and transcriptome datasets generated for 201 post-mortem human retina samples from individuals of European ancestry, with a median age of 71 (IQR=67-77) and without signs of late-stage AMD [20]. These data were derived from short-read whole genome sequencing (WGS; *n*=201) that achieved an average genome-wide coverage of 36x (IQR=30-41x), and polyA-enriched bulk short-read RNA sequencing for neurosensory retina (NSR; *n*=183; 139M paired-end reads, on average) and retinal pigment epithelium (RPE; *n*=176; 62M paired-end reads, on average). Utilising single-nucleotide variants (SNVs) detected from WGS with an internal minor allele frequency >1% and within 1Mb of transcription start sites, we generated significant predictive gene expression models for 22,036 and 22,034 genes in NSR and RPE, respectively. Next, we performed a discovery AMD-TWAS through integration with findings from a recent AMD-GWAS, including 57,290 AMD cases and 324,430 controls of European ancestry [8]. These analyses were restricted to 20,462 genes in NSR tissue and 20,463 genes in RPE tissue with predicted gene expression models and with appropriate SNV coverage from the GWAS. Through this approach, we identified 115 genes in NSR with significant AMD associations (***Figure 1a***; Bonferroni-corrected significance threshold, *p* < 2.44 × 10^−6^), 34 of which have not previously been associated with AMD-risk (***Supplementary Table 1 and 2***). Similarly, 104 genes were identified with significant AMD associations in RPE (***Figure 1b***), with 32 of these genes not previously associated with AMD-risk (***Supplementary Table 1 and 3***). For genes with expression models available in both NSR and RPE (*n*=20,442), AMD-TWAS associations were largely consistent between the tissues (***Figure 1c***), with 88 genes showing significant AMD association in both tissues, of which 78 had the same direction of effect (68% of NSR and 75% of RPE significant associations). In both tissues, the most significantly associated gene with AMD-risk was *HTRA1-AS1 (HTRA1 and ARMS2 antisense RNA 1; ENSG00000285955*), a long non-coding RNA (lncRNA) at the chromosome 10 (*ARMS2/HTRA1*) AMD-risk locus (***Figure 1***).

**Figure 1.**
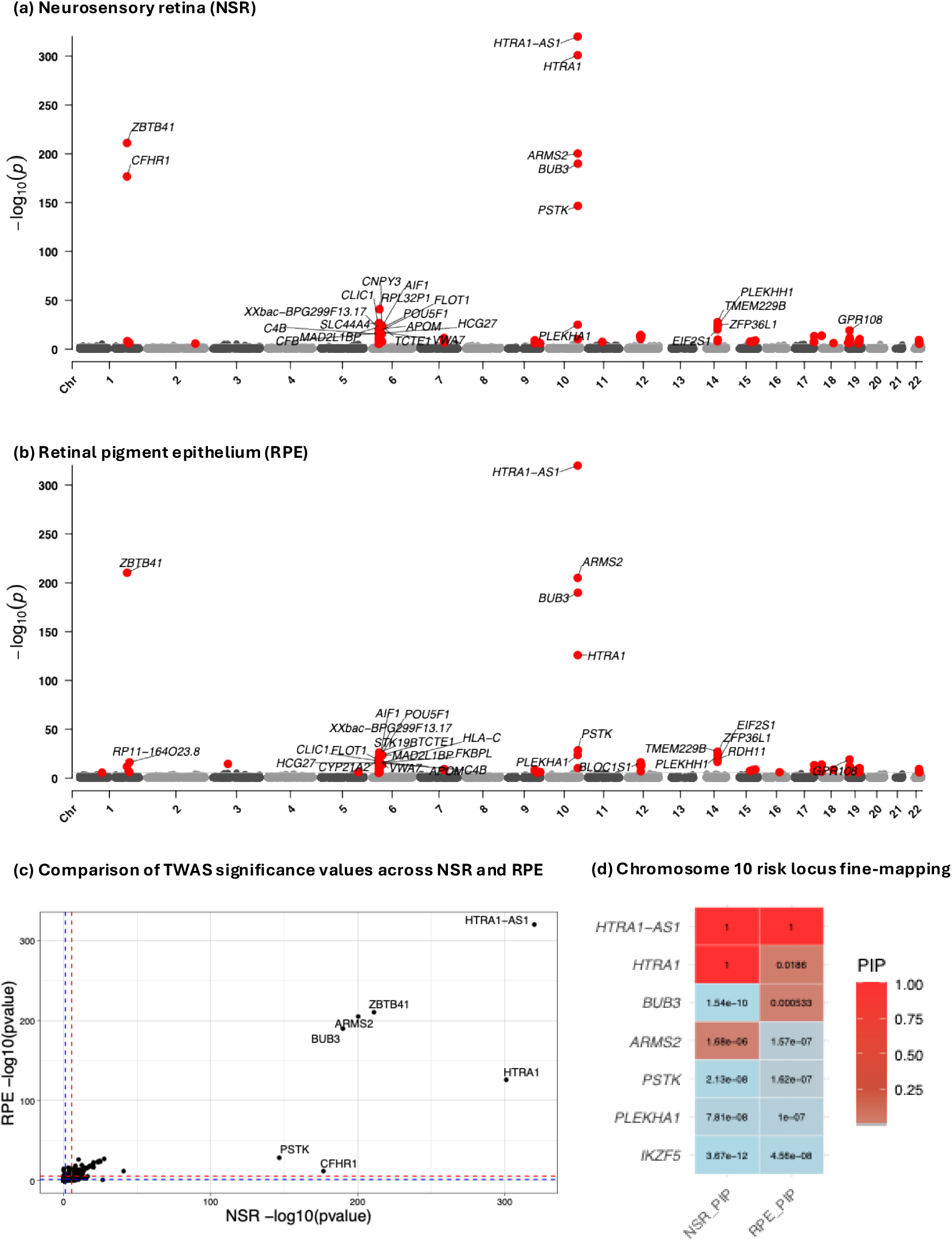
Discovery AMD-TWAS and fine-mapping. Integrating GWAS from 57,290 age-related macular degeneration (AMD) cases and 324,430 controls and short-read RNA sequencing from neurosensory retina (NSR, n=183) and retinal pigment epithelium (RPE, n=176) highlights *HTRA1-AS1* as a robust AMD-risk candidate gene across both tissues. *PIP*, posterior inclusion probability calculated by MA-FOCUS.

To assess reproducibility of these findings, we repeated the TWAS analyses with findings from an independent AMD-GWAS study, including 12,495 AMD cases and 461,686 controls of European ancestry [12]. In NSR, 74 of the 115 genes significantly associated in the discovery analysis were also nominally associated (*p*<0.05) with AMD in the validation analyses, with consistent effect direction for 71 of these genes (***Supplementary Table 2***). Similarly, 72 of the 104 gene signals from RPE attained nominal significance for association with AMD in the validation analyses with the same direction of effect (***Supplementary Table 3***). Of note, at the chromosome 10 AMD-risk locus, *PLEKHA1* was the only gene to be identified as significant in both discovery and validation cohorts but with opposite direction of effects, and *HTRA1-AS1* was the most significantly AMD-associated gene in the validation TWAS analyses across both NSR and RPE (***Supplementary Tables 2 & 3***).

Our TWAS analyses identified genomic regions harbouring multiple significantly associated AMD-risk genes, including the chromosomes 10 locus. Such patterns of association could reflect multiple genes with independent effects on AMD or LD between SNVs included in the predictive gene expression models [16]. To further interrogate these regions and identify the genes most likely to be driving AMD-risk we performed fine-mapping with Multi-Ancestry Fine-mapping of CaUsal gene Sets (MA-FOCUS) [22, 23] and applied a posterior inclusion probability (PIP) threshold of >0.8. Fine-mapping was performed across 16 regions for NSR and 16 regions for RPE identified with multiple gene-level associations in the discovery AMD-TWAS. At complex loci, including a region on chromosome 6 with multiple HLA genes, fine-mapping resolved the number of associated AMD genes (PIP >0.8) from 45 to 3 in NSR (*SLC44A4, POU5F1, HCG27*; ***Supplementary Table 4***) and from 39 to 3 in RPE (*POU5F1, HCG27* and *STK198*; ***Supplementary Table 5***). At the chromosome 10 AMD-risk locus, significant gene-level AMD associations from the discovery AMD-TWAS (including *ARMS2, HTRA1 and PLEKHA1)* were resolved to 3 genes in NSR and 3 genes in RPE, with *HTRA1-AS1* the only significantly associated gene after fine-mapping across both tissues (***Figure 1d***; PIP=1).

To further investigate factors driving the *HTRA1-AS1* significant AMD-association from TWAS and fine-mapping approaches, we assessed the location and the impact of informative genetic variants on the level of expression of genes in the chromosome 10 region (***Figure 2a***). We observed that *HTRA1-AS1* is more highly expressed in bulk NSR than RPE but is modestly expressed compared to other chromosome 10 risk genes (***Figure 2b***), with median transcript-per-million values of 1.3 (IQR=0.9-1.8) in NSR and 0.5 (IQR=0.3-0.7) in RPE. We note that the originally characterised GWAS variant impacting the suspected promoter region of *HTRA1* intersects the first exonic region of *HTRA1-AS1* (rs11200638; ***Figure 2a***) and has opposing associations on the expression of these genes in the presence of the increased AMD-risk allele (***Figure 2c***). Moreover, we show that rs11200638 AMD-risk genotypes in NSR without signs of late-stage of AMD drastically alter the co-expression relationship between *HTRA1-AS1* and *HTRA1* (***Figure 2d***), suggesting that interaction and/or pathway regulation is disrupted in the presence of AMD-risk genotypes. Similar, but less pronounced, co-expression relationships were observed between *HTRA1-AS1* and *PLEKHA1* in NSR, but not with *ARMS2* (***Supplementary figure 1***). Trends observed in RPE for homozygous risk allele genotypes, although of interest, are likely driven by a small number of significant outliers (***Supplementary figure 2***).

**Figure 2.**
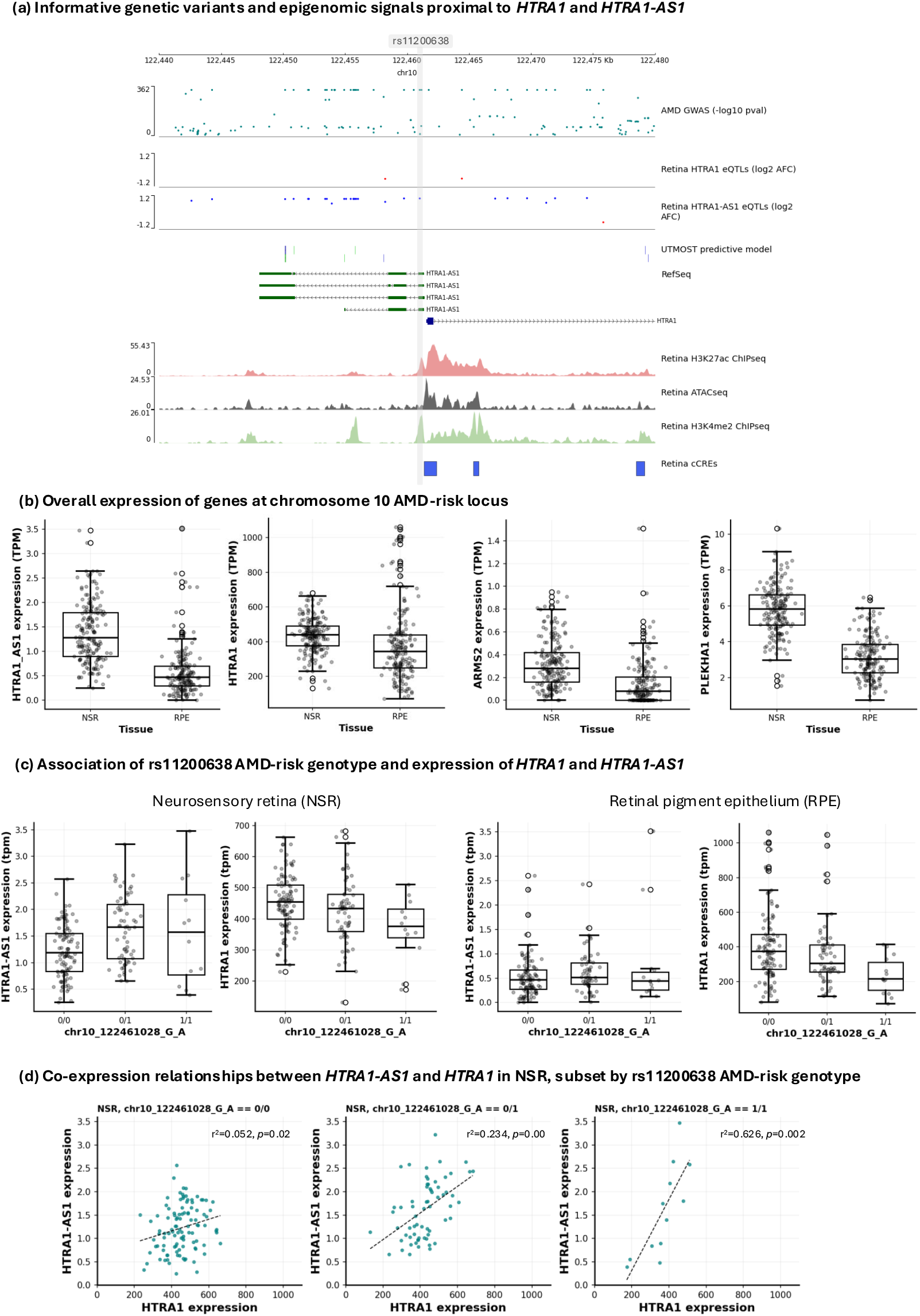
Genomic, transcriptomic and epigenomic composition of the chromosome 10 AMD-risk locus. Colours of variant locations in (a) indicate direction of effect (eQTLs) and gene assigned to (UTMOST predictive model).

Taken together, these data identify a new robust association between a lncRNA gene expressed in the adult human retina and AMD-risk, suggesting that genetic variation which dysregulates *HTRA1-AS1* expression is a critical component of AMD-risk. The directional effect of the TWAS and fine-mapping results performed in donor eyes without signs of late-stage AMD suggests that increased expression of *HTRA1-AS1* increases AMD-risk. Our findings are supported by recent preprints reporting that *HTRA1-AS1* has significantly dysregulated expression in AMD retinas compared to non-AMD donor retinas [24], is expressed in key retinal cell types and is associated with genetic risk of forming subretinal drusenoid deposits [25], a hallmark feature of AMD and risk of progression to late-stage AMD [26]. Molecular and cellular biology influenced by lncRNAs is yet to be fully characterised, although there are known roles in mature messenger RNA stability, chromatin function and regulation of co-expressed protein-coding genes [27-29]. Further understanding of the interaction between *HTRA1-AS1* and other genes in the chromosome 10 AMD-risk locus, including *ARMS2* and *HTRA1*, is an exciting opportunity to unpick the biological complexity driving early stages of AMD pathogenesis, with the potential to impact millions of individuals with increased risk of visual impairment and legal blindness. Our findings explain discrepancies in functional experiments where variant and protein-coding genes are assessed directly without this essential wider regulatory context, e.g. those reported for *HTRA1* [30, 31], and encourages revaluation of GWAS findings alongside moderately and co-expressed lncRNAs to further identify protein-coding genes that are implicated in disease risk due to their interaction with non-coding RNA genes.

In summary, we build on recent advances in the understanding of the role of non-coding genes [32, 33] and non-coding genomic regions [34-36] in visual impairment conditions impacting the retina, through identification that dysregulation of a lncRNA, *HTRA1-AS1*, is a key driver of AMD-risk at the well-established chromosome 10 risk locus.

## Methods

### Consent, ethical approval and data availability

This study utilises data that is publicly available. Whole genome sequencing and RNA sequencing was developed utilising donor eye tissues obtained from the Manchester Eye Tissue Repository, an ethically approved Research Tissue Bank (UK NHS Health Research Authority, 15/NW/0932), and is available through the European-Genome phenome Archive (EGA; Study ID: EGAS50000001443; Dataset: EGAD50000002082). GWAS statistics were downloaded following data deposits from the original papers [8, 12].

### Training a predicted gene expression model for adult human retinal tissue

To perform the Transcription-Wide Association Study (TWAS), we trained cross-tissue gene models using the unified test for molecular signatures (UTMOST) statistical framework [37]. We utilised genotype data and normalized gene expression data from bulk poly-A enriched RNA sequencing from adult human neurosensory retina (NSR) and retinal pigment epithelium (RPE). These data were developed and benchmarked as described elsewhere [20] and included whole genome sequencing for 201 individuals with paired RNA sequencing for NSR (n=183) and RPE (n=176). DNA and RNA sequencing was performed with standard library preparation procedures and paired-end sequencing using Illumina chemistry [20]. Whole genome sequencing datasets were processed with DRAGEN v4.0.3 and RNA sequencing datasets processed using the Genotype-Tissue Expression Consortium v8 pipeline [38], with transcripts characterised in Gencode v38. Expression quantitative trait loci were characterised with TensorQTL [39]. Gene expression quantification from RSEM [40] was restricted to genes with >0.1 transcripts-per-million and ≥6 reads in at least 20% of samples, and normalised using trimmed mean of M values in *edgeR* [41] and an inverse normal transformation. Training of predicted gene expression models in UTMOST included 55,253 autosomal genes, where for each gene, we considered only single nucleotide variants (SNVs) mapping within 1 Mb of the transcription start site and excluded SNVs with ambiguous alleles (AT or GC) or with internal minor allele frequency <1%, using default settings [21].

### Discovery and validation TWAS

Using the generated tissue-specific gene expression models, we performed TWAS by applying the UTMOST method to two recent GWAS of age-related macular degeneration (AMD). The discovery GWAS dataset included 57,290 AMD cases and 324,430 controls of European ancestry, aggregated from multiple large cohorts including the Million Veteran Program (MVP, tranches 1–3), the International AMD Genomics Consortium (IAMDGC), the Genetic Epidemiology Research on Aging (GERA) cohort, UK Biobank (UKBB), and Genentech’s GA and CNV datasets [8]. In the MVP cohort, AMD status was determined using ICD-9/ICD-10 codes from electronic health records, while other cohorts used their own criteria for defining cases and controls. To assess the reproducibility and robustness of our findings, we also performed an independent validation TWAS using datasets from the FinnGen release R12 [12]. This dataset includes 12,495 AMD cases (encompassing both dry and wet AMD) and 461,686 controls, with case definitions based on standardized FinnGen phenotyping. Novel associations between gene and AMD were identified through comparison of loci identified in previously published studies [7, 8, 10, 17, 42] and through assessment in data reported in the NHGRI-EBI GWAS catalog [43] and the Ocular Knowledge Portal (ocular.hugeamp.org).

### Fine-mapping of AMD-TWAS results

Fine-mapping of gene–trait association signals from the discovery TWAS was performed using Multi-Ancestry Fine-mapping of CaUsal gene Sets (MA-FOCUS) [44, 45], a gene-level statistical method that identifies likely causal genes through modelling the correlation structure induced by linkage disequilibrium and the expression prediction weights utilized in TWAS. To define input regions for fine-mapping, we selected all genes with transcriptome-wide significant associations with AMD (p < 2.44 × 10^−6^). For each significant gene, a genomic window spanning ±1 Mb from the transcription start site was defined, corresponding to the region from which SNVs were originally used to train the gene expression prediction model. When the windows of two or more significant genes overlapped, they were merged into a single non-redundant region to jointly analyse shared signals. MA-FOCUS was applied to each resulting region, including those containing multiple significant genes.

### Genomic, gene expression and epigenomic trends at chromosome 10 AMD-risk locus

All gene expression statistical trends and analysis were conducted in python and R, using standard outputs of the GTEx consortium v8 pipeline [38], and with datasets available through Gencode (v49) or previously published [46].

## Acknowledgements

We express our sincere thanks to the donors, and their families, for enabling this research. This research was supported by the Macular Society (United Kingdom), Fight For Sight, the UK Medical Research Council (MR/V020749/1) and the NIHR Manchester Biomedical Research Centre (NIHR203308). of Health and Social Care. The authors acknowledge the assistance of Research IT Support, the use of the Computational Shared Facility and the Genomic Technologies Core Facility at The University of Manchester. The views expressed in this article are those of the authors and do not necessarily represent those of the UK National Health Service, the UK National Institute for Health Research, or the UK Department of Health and Social Care. The authors thank Million Veteran Program (MVP) staff, researchers, and volunteers, who have contributed to MVP, and especially participants who previously served their country in the military and now generously agreed to enroll in the study. (See https://www.research.va.gov/mvp/ for more details). This research is based on data from the Million Veteran Program, Office of Research and Development, Veterans Health Administration, and was supported by the Veterans Administration (VA) Million Veteran Program (MVP) award #000. The authors acknowledge the participants and investigators of the FinnGen study.

---

**Supplementary Figure 1.**
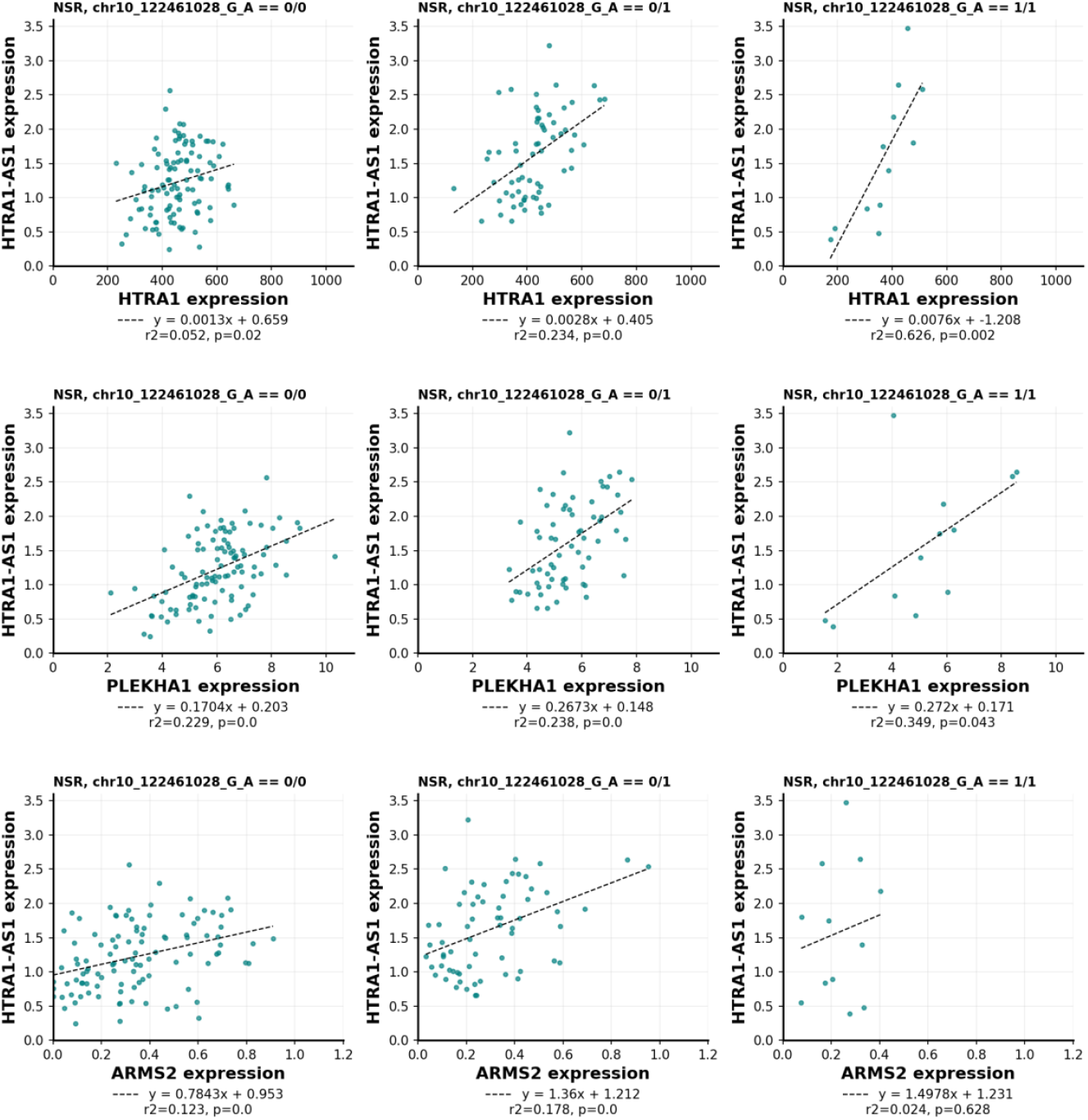
Co-expression relationships between *HTRA1-AS1* and protein-coding genes at the chromosome 10 AMD-risk locus for neurosensory retina samples (n=183).

**Supplementary Figure 2.**
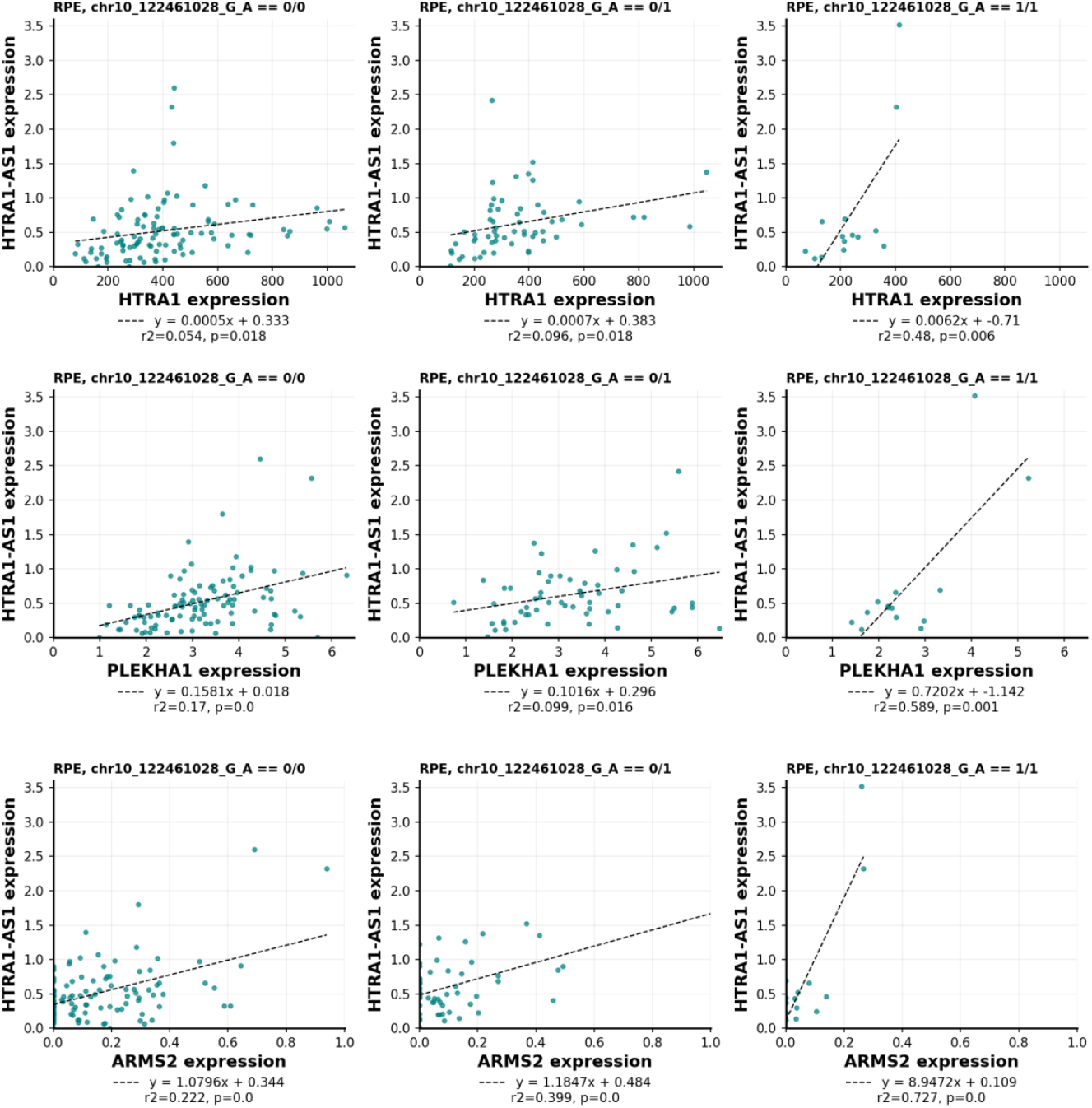
Co-expression relationships between *HTRA1-AS1* and protein-coding genes at the chromosome 10 AMD-risk locus for retinal pigment epithelium samples (n=176).

